# White matter microstructural changes in post-traumatic headache: A diffusion tensor imaging (DTI) study

**DOI:** 10.1101/2024.08.05.24310944

**Authors:** Ho-Ching Yang, Tyler Nguyen, Kelly M. Naugle, Fletcher A. White, Yu-Chien Wu

## Abstract

**Introduction:** Post-traumatic headache (PTH) is a common consequence of mild traumatic brain injury (mTBI) that can severely impact an individual’s quality of life and rehabilitation. However, the underlying neuropathogenesis mechanisms contributing to PTH are still poorly understood. This study utilized diffusion tensor imaging (DTI) to detect microstructural alterations in the brains of mTBI participants with or at risk of developing PTH.

**Method:** This study investigated associations between DTI metrics 1-month postinjury and pain sensitivity, as well as psychological assessments 6-months postinjury to identify differences between mTBI (n = 12) and healthy controls (HC; n = 10). MRI scans, including T1-weighted anatomical imaging and DTI were acquired at 1-month postinjury. Pain sensitivity assays included quantitative sensory testing and psychological assessment questionnaires at 1-month and 6-months postinjury.

**Results:** Significant aberrations of mean axial diffusivity in the forceps major were observed in mTBI relative to HCs at 1-month postinjury (p =0.02). Within the mTBI group, DTI metrics at 1-month postinjury were significantly associated (p’s < 0.05) with pain-related measures and psychological outcomes at 6-month postinjury in several white matter tracts (right sagittal stratum, left anterior thalamic radiation, left corticospinal tract, left insula, left superior longitudinal fasciculus). Notably, the associations between DTI metrics at 1-month postinjury and pain-related measures at 6-month postinjury showed significant group differences in the right sagittal stratum (p’s < 0.01), white matter tract in left insula (p < 0.04), and left superior longitudinal fasciculus (p’s < 0.05).

**Conclusion:** This study suggests that “Post-Traumatic Stress Disorder for DSM-5” and “Center for Epidemiological Studies-Depression Scale” are the most sensitive psychological measures to early microstructural changes after mTBI, and that the DTI metrics are predictive of pain and psychological measures in mTBI. Together, these results suggest that white matter microstructure plays an important role in the PTH following mTBI.

## INTRODUCTION

Traumatic brain injury (TBI) is a major public health problem in the United States [1]. The overwhelming majority (∼80%) of TBIs are categorized as “mild”, a term that may be misleading, as some individuals recover more slowly than anticipated and a minority develop chronic neurobehavioral sequelae [2]. Individuals sustaining mTBI have a wide spectrum of clinical signs and symptoms, including headache, cognitive impairment, psychological distress, and somatic symptoms. Post-traumatic headache (PTH) is one of the worst, most prominent, and longest-lasting consequences of mTBI. It is defined as a secondary headache that develops within seven days following the injury [3]. PTH is regarded as chronic when it continues for more than three months, with 18-22% lasting more than one year [4, 5] and approximately 2-5% lifetime prevalence [6]. Persistent PTH severely impacts quality of life and sabotages rehabilitative efforts. Therefore, effective intervention and treatment will benefit millions of affected individuals, if the underlying neuropathogenesis mechanisms in persistent PTH can be clarified by identifying reliable biomarkers.

Despite the high prevalence and persistence, the underlying neuropathogenesis mechanisms in persistent PTH are still poorly understood. Persistent PTH may arise from peripheral origin triggering neurogenic inflammation or central origin damaging the pain-related pathways after the injury [5]. The unique origins of persistent PTH suggest therapeutic strategies that differ from those for migraines and primary headaches.

Diffuse axonal injury is generally believed to be the initial neuropathology associated with mTBI [7–9]. A few studies report evidence of diffuse axonal injury in *postmortem* pathological examination of human mTBI. These papers suggest severing of nerve fibers without hemorrhage [10], the presence of amyloid precursor protein indicating axonal damage [11], and polarized macrophages in white matter [12]. Additionally, evidence of axonal injury in conjunction with chronic traumatic encephalopathy has been demonstrated in football players exposed to repetitive head impacts and/or concussion [13].

Nevertheless, this microscopic white-matter injury has been more difficult to detect in human mTBI with *in vivo* approaches. Neuroimaging may provide a significant advance for quantitatively detecting and characterizing the mechanisms of brain changes in PTH. With mostly negative findings, *conventional* diagnostic MRI and CT exams, however, are unable to provide useful diagnostic and prognostic utility. Diffusion MRI may be more sensitive than conventional imaging in detecting white-matter changes in mTBI [14–16].

Among different diffusion MRI approaches, the classic diffusion tensor imaging (DTI) has been widely used in white-matter diseases and was found to have adequate diagnostic sensitivity to microstructural changes in the brain after mTBI [17–19]. Specifically, DTI produces two summary metrics, fractional anisotropy (FA) and mean diffusivity (MD), and two orthogonal metrics, axial diffusivity (AD) and radial diffusivity (RD). ***FA***, the variance of the three eigenvalues of a diffusion tensor, indicates how directional the diffusion is and reflects the coherence of underlying tissue or fiber organization. ***MD***, the mean of the three eigenvalues of a diffusion tensor, describes the general freedom of diffusion in the tissue regardless of the directionality. FA and MD have been the main focus in DTI studies of mTBI [18] and PTH [20]. Although less-discussed, AD and RD may provide a higher level of biological specificity than FA and MD. ***AD***, the largest eigenvalue of the tensor, describes the longitudinal diffusivity along the white-matter fiber direction, while ***RD***, the average of median and minor eigenvalue, describes the transverse diffusivity perpendicular to the fiber.

In this pilot study, we will include all four metrics to interpret DTI results and to better characterize microstructural alterations in the brain with PTH. We studied (1) group differences (mTBI and control) at 1-month postinjury, (2) associations between early DTI metrics at 1-month postinjury and later pain and psychological related assessments at 6-month postinjury to detect predictivity of DTI metrics in mTBI, and (3) for those significant within group associations, if there are group differences in such prediction power.

## METHODS

### Participants

Details on the recruitment, diagnoses of mTBI, inclusion, and exclusion criteria can be found in our previous publication [21, 22]. Briefly, mTBI participants were recruited from Level 1 trauma centers within hospitals located in the Indianapolis area. Study coordinators screened the electronic medical records of patients with a head injury that were admitted to the Emergency Department and identified whether patients met the inclusion and exclusion criteria. A mild TBI diagnosis was also confirmed by the attending Emergency Department Physician according to the criteria recommended by the World Health Organization Task Force [23]. Then, the identification and contact information of potentially eligible mTBI patients were put into a secure database. The research staff would then call or email the potential participant to determine whether the individual was interested and eligible for the study. For individuals interested and potentially eligible, the first study visit was scheduled.

Inclusion criteria for the mTBI participants included: 1) a Glasgow Coma Scale score between 13 and 15 when examined at the emergency center, 2) no abnormal findings on a computed tomography scan of the brain to exclude secondary disorders, 3) the presence of one or more of the following: confusion or disorientation, post-traumatic amnesia for less than 24 h, or a loss of consciousness for less than 30 min, and 4) between the ages of 18-65 years old at the time of enrollment. Exclusion criteria included: 1) chronic cardiovascular disease or uncontrolled hypertension, 2) metabolic disease, renal disease, hepatic disease, or transplant conditions, 3) neurological disease, 4) serious psychiatric conditions or hospitalization within the preceding year for psychiatric illness, 5) chronic headaches before the head injury, 6) current involvement in litigation, 7) chronic use of narcotics, 8) fracture or polytrauma at the time of head injury, 9) metallic foreign bodies in the brain or eye, metal braces in the mouth, or cardiac pacemakers, 10) significant cognitive impairment as indicated the Mini Mental Status Examination (MMSE) score of ≤ 23, and 11) pregnant women as determined by a FDA-approved commercial urine pregnancy test. All mTBI participants received pain and psychological assessments at 1-month and 6-months postinjury. A subset of the mTBI participants received MRI scans at 1-month postinjury and was included in this study to study the relationship and predictivity of neuroimaging and later pain-related outcomes. Thirteen mTBI participants received neuroimaging at 1-month postinjury. One participant’s MRI data was excluded due to an extended artifact from the dental implant. Thus, the final sample size for the mTBI group was 12.

Ten age matched control participants with no TBI history were recruited from the community with the same exclusion criteria. The controls completed two study sessions separated by 5-6 months and received the same pain and psychological assessments and neuroimaging. All participants reviewed and signed a written informed consent form approved by the Indiana University Institutional review board (IRB).

### Pain and psychological assessments

#### Quantitative sensory tests

We used quantitative sensory tests (QST) to measure pain sensitivity of the head and endogenous pain modulation. These tests included temporal summation of pain (TS) as an indirect method of assessing hyperexcitability of the CNS [24], pressure pain thresholds measuring trigeminal sensitization (PPTs) [25], and conditioned pain modulation (CPM) measuring endogenous pain inhibition [26, 27]). Participants were first familiarized with each QST test and taught the 0-100 numeric pain rating scale. The TS test was performed first, followed by PPTs of the head, and then the CPM test. At least 10 minutes separated each QST test. All participants were asked to refrain from pain-relief medication and consuming caffeine on the day of testing before each session.

##### Mechanical Temporal Summation

TS was tested on the middle of the forehead and the back of the hand using a Von Frey filament (Touchtest Sensory Evaluator 6.65) calibrated to bend at 300g of pressure. First, a single pinprick with the filament was applied to the body site. Participants verbally rated pain intensity of the single pinprick on the 0 to 100 pain scale. Then, a series of 10 pinpricks with the same filament was applied to the same body site within an area of 1 cm^2^ and at a rate of 1 prick per second. Immediately after the 10 pinpricks, participants rated their greatest pain experience during the 10 pinprick stimuli using the 0 to 100 scale. This TS procedure was administered two times at each body site with 60 seconds between trials. The two trials at each body site were averaged for a single TS forehead score and a single TS hand score.

##### Pressure Pain Thresholds of the Head

PPTs were assessed on the head by applying a clinical-grade pressure algometer (AlgoMed, Medoc Advanced Medical Systems, Durham, NC, USA) with a 1.0 cm^2^ probe against the skin on five sites of the head and neck including: the middle of the forehead, the left temple, the parietal area (top of head), the posterior neck/C2, and the left trapezius. Pressure from the algometer was applied at a slow constant rate of pressure (30kPA/s). Participants pressed a button when they first experienced pain caused by the pressure algometer. The algometer was then removed and the pressure was recorded. Two PPT trials were administered at each body site with 20-seconds separating each trial. The PPT scores for all trials and body sites were averaged for a single PPT score, which was used for data analysis.

##### Conditioned Pain Modulation

CPM is the most common test of endogenous pain inhibition in humans and utilizes a “pain-inhibition-by-pain” model. CPM assesses whether pain produced by a test stimulus is reduced by a second painful conditioning stimulus applied to a remote body site. [28, 29] For the CPM protocol used in the current study, PPTs (test stimulus) were administered on the left volar forearm before (pretest) and immediately after (posttest) the submersion of the right hand in a cold water bath (conditioning stimulus). The test stimulus of PPTs was administered using the same procedures as described above, except at a different body location. Two PPT trials were administered for each pre– and post-conditioning test. For the conditioning stimulus, the right hand was placed in a cold water bath maintained at 10°C for 1 minute. To measure the magnitude of CPM, a percent change score was calculated for the test stimulus with the following formula: [(post-test PPT trial score – pre-test PPT trial score)/ pre-test PPT trial score] *100. A positive percent change score indicated that PPTs increased following the conditioning stimulus, indicative of pain inhibition.

##### Psychological and Clinical Pain assessments

Head Pain and psychological measures were collected via validated questionnaires as described below.

##### Headache Survey

Participants completed a Headache Survey that has been used in prior research of post-traumatic headache [30, 31]. The survey included questions regarding the intensity, frequency, and duration of headaches, medication use and other treatments, headache triggers, preinjury history of headache, and symptoms of ongoing headache. The average headache pain intensity rating was used in data analysis. Participants rated on a 0 to 10 numeric rating scale (NRS) the average pain intensity of their headaches during the past week, with 0 indicating no headaches at all and 10 indicating the worst pain possible.

##### McGill Pain Questionnaire (MPQ)

The MPQ provides a quantitative evaluation of a person’s pain experience with separate sensory, affective, evaluative, and miscellaneous dimensions [32]. The MPQ is composed of a list of 78 words categorized into 20 groups of words representing the four dimensions. From each group of words, respondents choose the word that best describes their experience of pain. Participants completed the MPQ to describe their present head pain. For data analysis, we calculated the pain rating index (PRI), which sums the rank values of the words chosen by the participant from the word list. The MPQ has been used extensively in clinical and research settings and has been validated within TBI populations [32, 33].

##### Defense and Veterans Pain Rating Scale (DVPRS)

THE DVPRS is a 0 to 10 scale in which participants rate their current pain, with 0 indicating “no pain” and 10 indicating “as bas as it could be, nothing else matters” [34].

##### TBI Quality of Life (TBI-QOL) Headache Pain Form

This questionnaire assesses the severity and impact of headaches over the last 7 days with 10 items.

##### Post-Traumatic Stress Disorder (PTSD) for DSM-5 (PCL-5)

The PCL-5 is a 20-item scale that assesses the DSM-5 symptoms of PTSD, and is used for diagnosis and as a severity measure. [35]

##### Pain Catastrophizing Scale (PCS)

The PCS includes 13 items that evaluate respondents’ emotional and cognitive responses to actual or anticipated pain, including rumination, magnification, and helplessness [36]. Higher scores indicate greater pain catastrophizing.

##### Center for Epidemiological Studies – Depression Scale (CES-D)

The CES-D includes 20-items that assess symptoms associated with depression. The total score ranges from 0 to 60, with higher scores indicative of increased depression. The CES-D is valid and reliable in the general and clinical populations [37].

#### Image Acquisition

The participants underwent MRI scans in Siemens MAGNETOM Biography mMR 3T scanner with a 20-channel head/neck coil. The MRI scans included T1-weighted anatomical imaging and diffusion-weighted imaging. The T1-weighted images were acquired by inversion recovery-prepared spoiled gradient-echo (IRSPGR) with TR/TE =1960/2.19 ms, flip angle = 10 deg, and 1.0X1.0X1.0*mm*^3^ voxels. Diffusion MRI was performed with a single-shot echo planar imaging sequence with a single 180-degree refocusing pulse. The diffusion-encoding scheme consisted of 64 directions at a b value of 1000 s/*mm*^2^ and 1 b0 (b value = 0 s/*mm*^2^). Other imaging parameters were filed-of-view = 2080 mm, 60 slices, voxel size = 2.0X2.0X2.0 *mm*^3^, TR/TE = 10900/79 ms, flip angle = 90 deg, in-plane GRAPPA acceleration factor = 2. For distortion correction, a dual-echo gradient echo sequence (GRE) was used to acquire the fieldmap with TR = 400 ms, TE = 4.92/7.38 ms, flip angle = 60 deg, and 3.0X3.0X3.0*mm*^3^voxels.

#### Diffusion MRI data analysis

The diffusion-weighted images were first denoised with the Marchenko-Pastur principal component analysis approach [38]. Gibbs ringing artefacts were removed from diffusion-weighted images using the method of local subvoxel-shifts (MRtrix3 mrdegibbs) [39]. With a dual-echo GRE field map and co-registered T1-weighted image, the diffusion-weighted images were then corrected for motion, eddy current artifacts, and static-field geometric distortion with the eddy_openmp command provided in the FMRIB Software Library (FSL) [40]. The FSL eddy command detects outliers’ slice by slice with Gaussian process predictions. B1 field inhomogeneity was also corrected (MRtrix3 dwibiascorrect) [41]. Four classic DTI metrics were calculated and registered to Montreal Neurological Institute space using Advanced Neuroimaging Tools nonlinear registration [42]. These metrics included fractional anisotropy (FA, describing the variance among the eigenvalues of the diffusion tensor), mean diffusivity (MD, describing the speed of water movement (i.e., diffusion) in the brain tissue regardless of the directionality), radial diffusivity (RD, describing diffusion perpendicular to the axonal axis), and axial diffusivity (AD, describing diffusion parallel to axons) An abnormal decrease in FA may indicate disorganization of axons or demyelination. An abnormal increase in MD may indicate destruction of the tissue microarchitectures such as axonal beading, cellular swelling, demyelination, or brain edema. An abnormal increase in RD indicates structural destruction perpendicular to the axons such as demyelination, while an abnormal increase in AD indicates structural destruction parallel to the axons such as destruction of cytoskeletons.

After the maps of the DTI metrics were computed and transformed to the standard Montreal Neurological Institute (MNI) space, tract-based spatial statistics (TBSS) provided by the FSL toolbox was applied to the diffusion scaler maps in the MNI space to create a whole-brain white matter skeleton [43]. White matter skeletond were extracted and segmented to 18 bilateral region-of-interests (ROIs) [43, 44], including the insular, cingulum cingulate gyrus, hippocampus, corticospinal tract, posterior thalamic radiation with optic radiation, sagittal stratum with inferior longitudinal fasciculus and inferior frontal occipital fasciculus, superior longitudinal fasciculus, anterior thalamic radiation, forceps major, and forceps minor (Figure 1). The MNI Structural Atlas was selected for the bilateral insula [45]; The other 16 ROIs were selected using JHU White-Matter Tractography Atlas [46].

**Figure 1.**
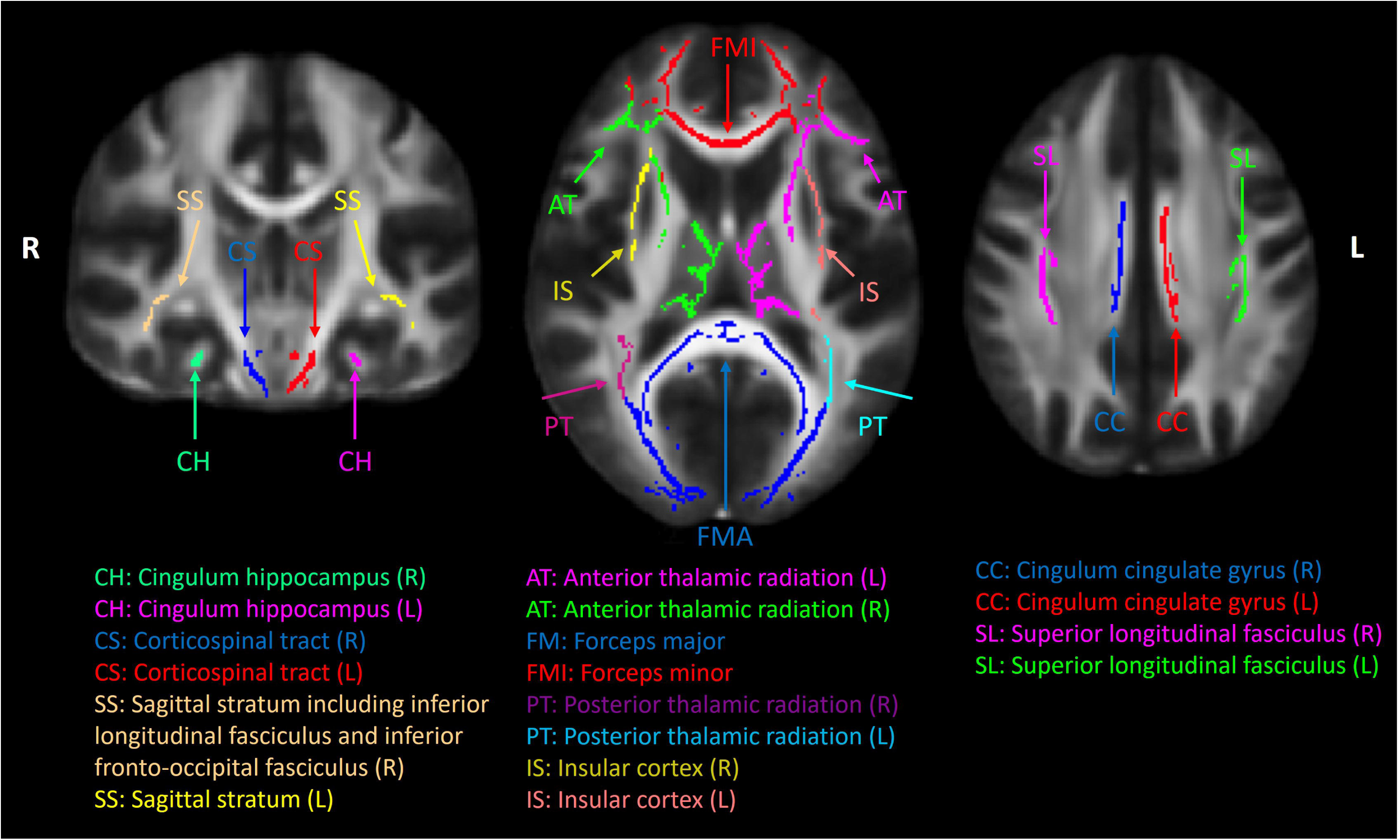
Cerebral white matter region of interests (ROIs). A common white matter skeleton was first generated by the tract-based spatial statics (TBSS) toolbox in FSL using normalized DTI factional anisotropy maps from all the subjects. The white matter skeleton was intercepted with Johns Hopkins University (JHU) white matter atlas and eighteen cerebral ROIs were used in this study.

#### Statistical analysis

The study utilized two sample T-test to examine group differences in continuous variables related to demographic and clinical outcome measures, as presented in table 1. Sex, as a categorical variable was analyzed by x^2^ tests to assess differences in sex compositions across groups. Generalized linear regression models were used to test 1) group differences in the DTI metrics between mTBI and control participants at 1-month, 2) associations of the early DTI metrics at 1 month with later outcome measures at 6 months to assess predictivity of DTI in mTBI, and 3) group differences in the predictivity of DTI for pain and psychological outcome measures. For the group differences in the DTI metrics, we used Model 1 controlling for age, sex, and the DVRS pain rating at the time of MRI scans (Eq. 1). For the predictivity evaluation in the mTBI group, we used Model 2 controlling for age, sex, and the DVRS scores (Eq. 2). For those significant associations in Model 2, we further tested if there are group differences in the predictivity of DTI using Model 3 (Eq. 3), which included an interaction term of ‘group*DTI’. Specifically, if β_3_ in Model 3 is significant (i.e., *p*_int_ < 0.05), there is significant group differences in the associations between outcomes_6-mo_ and DTI_1-mo_. When the interaction term is significant (i.e., *p*_int_ < 0.05), we performed post hoc correlation analyses in the control group using Model 2 and compare the scatter plots between the two groups. P-values less than 0.05 were deemed significant. Due to exploratory nature of this pilot study and small sample size, we did not adjust for multiple comparisons.

Model 1:

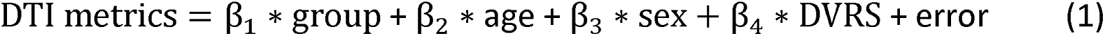

Model 2:

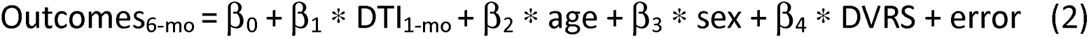

Model 3:

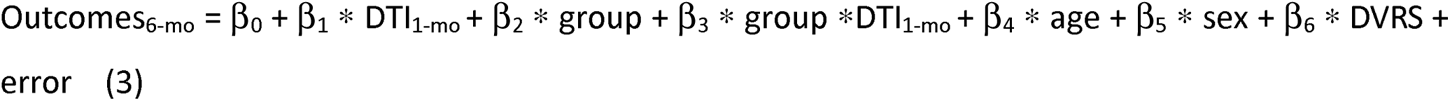

## RESULTS

The demographic and characteristics of the participants are listed in Table 1. There were no significant differences between the mTBI and control participants in age and sex. Compared to the controls, the mTBI participants had significantly higher psychological measures (i.e., PCL (*p* = 0.03) and CESD (*p* < 0.05)) at 1-month postinjury and higher scores in all the clinical pain measures (i.e., HA, MPQ, TBIQL, and DVRS) (p’s < 0.01) at 1-month and 6-month postinjury.

### Group differences in the DTI metrics

The mTBI participants exhibited significantly lower AD in the forceps major (*p* = 0.02) than the controls at 1-month postinjury (Figure 2). Other DTI metrics and white-matter fiber tracts did not differ significantly between groups.

**Figure 2.**
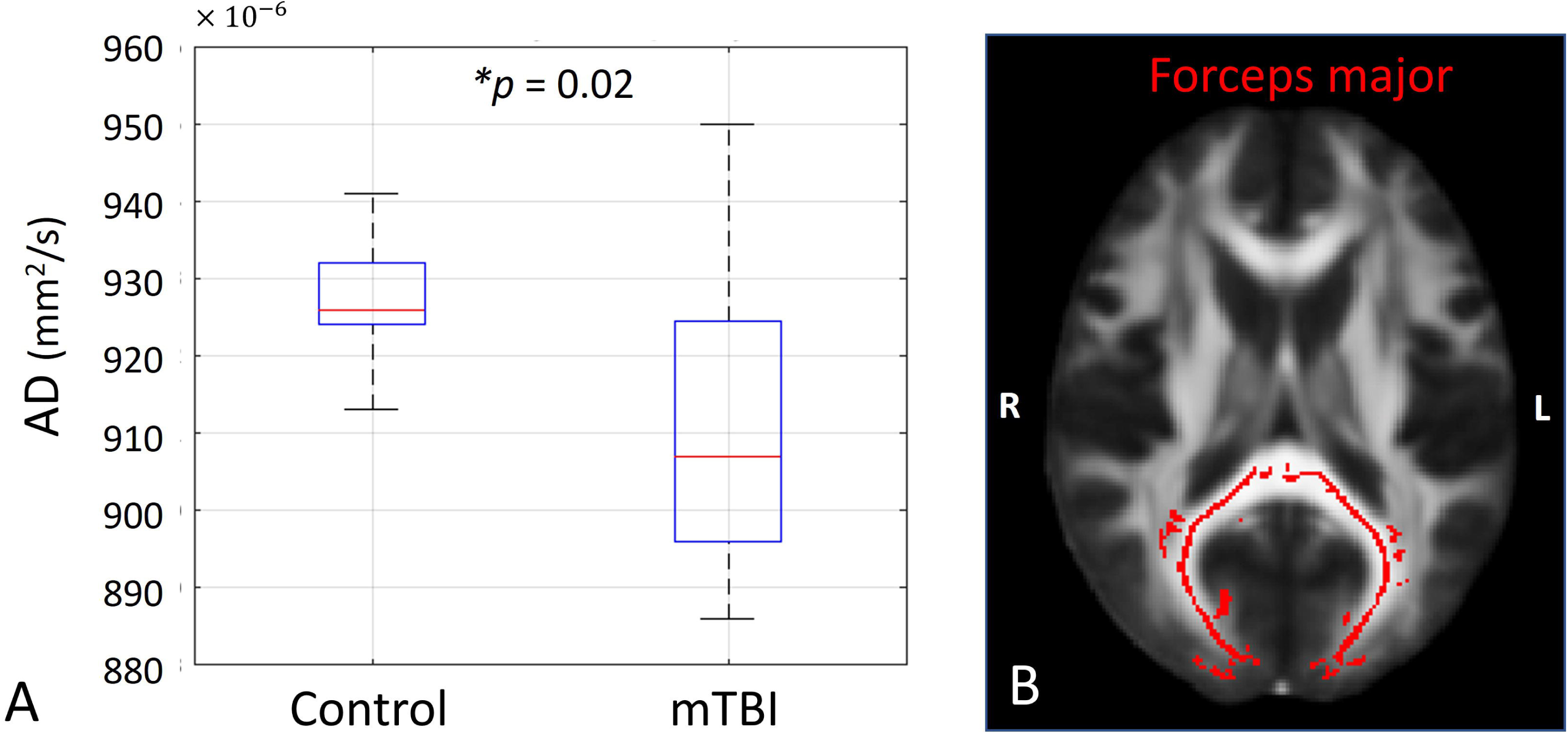
(A) Bar plot of DTI axial diffusivity (AD) between the mTBI participants and controls in the forceps major at 1-month postinjury. * denotes significant p value less than 0.05. (B) The forceps major fiber tract is highlighted in red. The image is presented in radiology orientation.

#### Association between the early DTI metrics and later pain and psychological related measures

Within the mTBI group, the DTI metrics at 1-month postinjury had significant associations with the pain-related measures at 6-month postinjury (Table 2). The QST TS of the head was predicted by DTI FA in the right sagittal stratum, including the inferior longitudinal fasciculus and inferior frontal occipital fasciculus, with a large correlation coefficient, r = –0.60 (*p* < 0.05). The QST PPT was predicted by DTI AD in the left anterior thalamic radiation and corticospinal tract with a moderate correlation coefficient, r > 0.20 (*p* < 0.05). Furthermore, QST CPM had a significant positive association with FA (r = 0.83, *p* < 0.05).

The DTI metrics also predicted later psychological outcomes (Table 2, middle). PCL and CESD were predicted by both FA (negative association) and RD (positive association) in the right sagittal stratum with large correlation coefficients (|r| > 0.60, *p* < 0.05). They were also predicted by MD and RD in left superior longitudinal fasciculus (positive association, r > 0.73). PCS was predicted by MD in the left insular and AD in the left corticospinal tract (positive association, r > 0.68, *p* < 0.05).

Regarding the clinical pain measures, the MPQ pain scale at 6-month also increased with elevated RD in the left Corticospinal tract (r = 0.67, *p* < 0.05) at 1-month postinjury. All the above associations had a median goodness of fit (R2) of 0.75 (0.67 – 0.94).

#### Group differences in the associations between DTI and pain-related measures

The associations between the early DTI metrics and later pain-related measures had significant group differences in three cerebral white-matter tracts, including the left insula, right sagittal stratum, and left superior longitudinal fasciculus (Table 3 and Figures 3A, 4A, and 5A). While the mTBI group had significant associations indicating the predictivity of the early DTI metrics and later pain and psychological outcomes (Table 3), the control group did not have any significant associations (*p*con > 0.05, Table 3). Overall, PCL and CESD are the most sensitive psychological measures to early microstructural changes after mTBI. The directionality of the associations can be appreciated by the scatter plots in Figures 3-5, where worse psychological scores in the mTBI group were associated with and could be predicted by elevated diffusivities (MD and RD) and reduced tissue micro-organization (FA).

**Figure 3.**
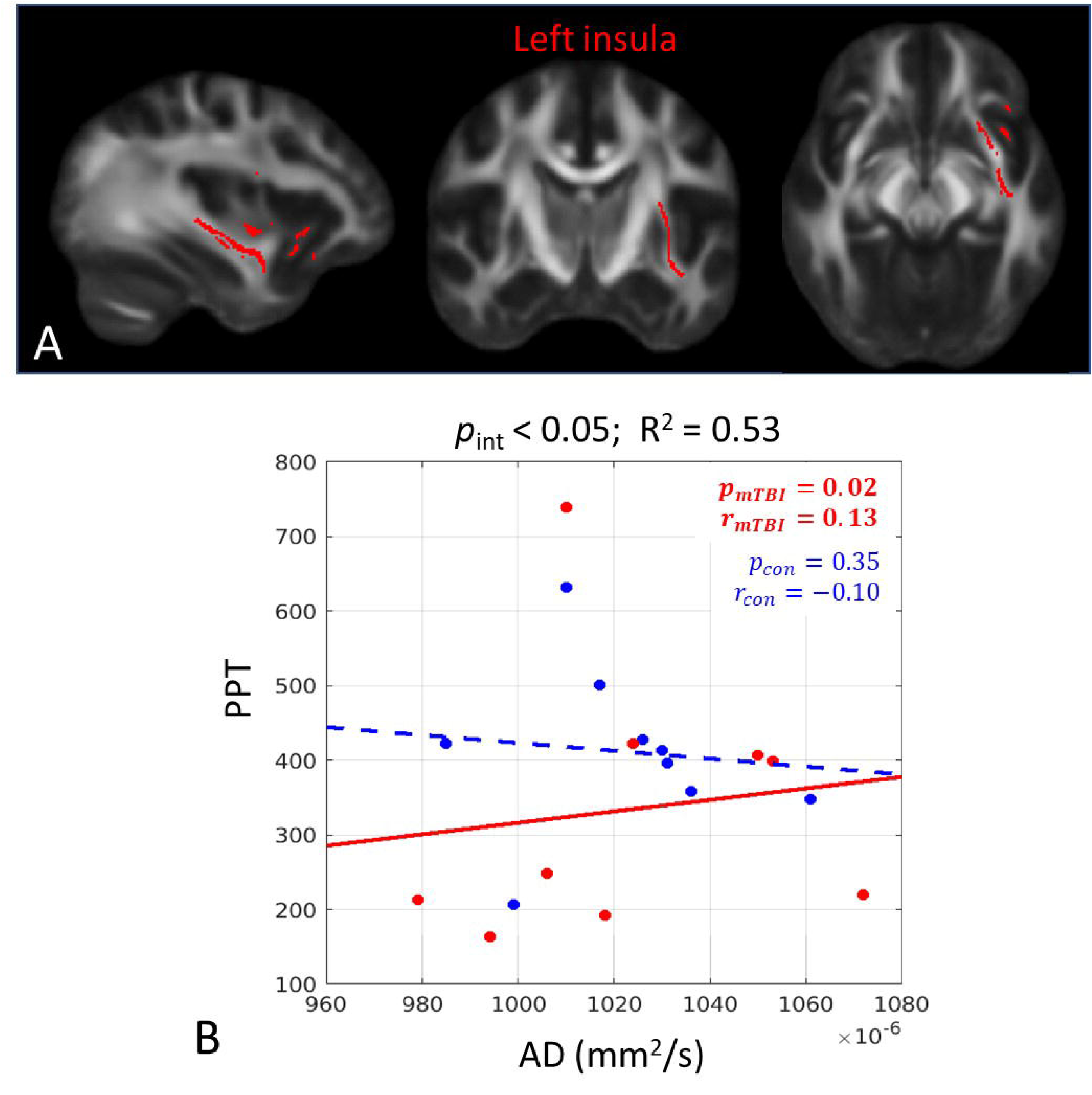
In the left insula, there was a significant group difference in the associations between axial diffusivity (AD) at 1-month postinjury and the pressure pain threshold of the trigeminal sensitization (PPT) at 6-month postinjury. (A) The significant white matter tract in the left insula is shown in red. (B) Scatter plots with post hoc correlation results. The mTBI group had a significant positive association (red solid line) while the control group did not (blue dashed line). *p*_int_ = significance level of the group interaction term in Eq. 3; R^2^ = overall coefficient of determination for goodness of fit for Model 3; *p*_mTBI_ = significance level of the association for the mTBI group; r_mTBI_ = correlation coefficient for the mTBI group; *p*_con_ = significance level of the association for the control group; r_con_ = correlation coefficient for the control group Bold numbers indicate statistical significance with p < 0.05.

**Figure 4.**
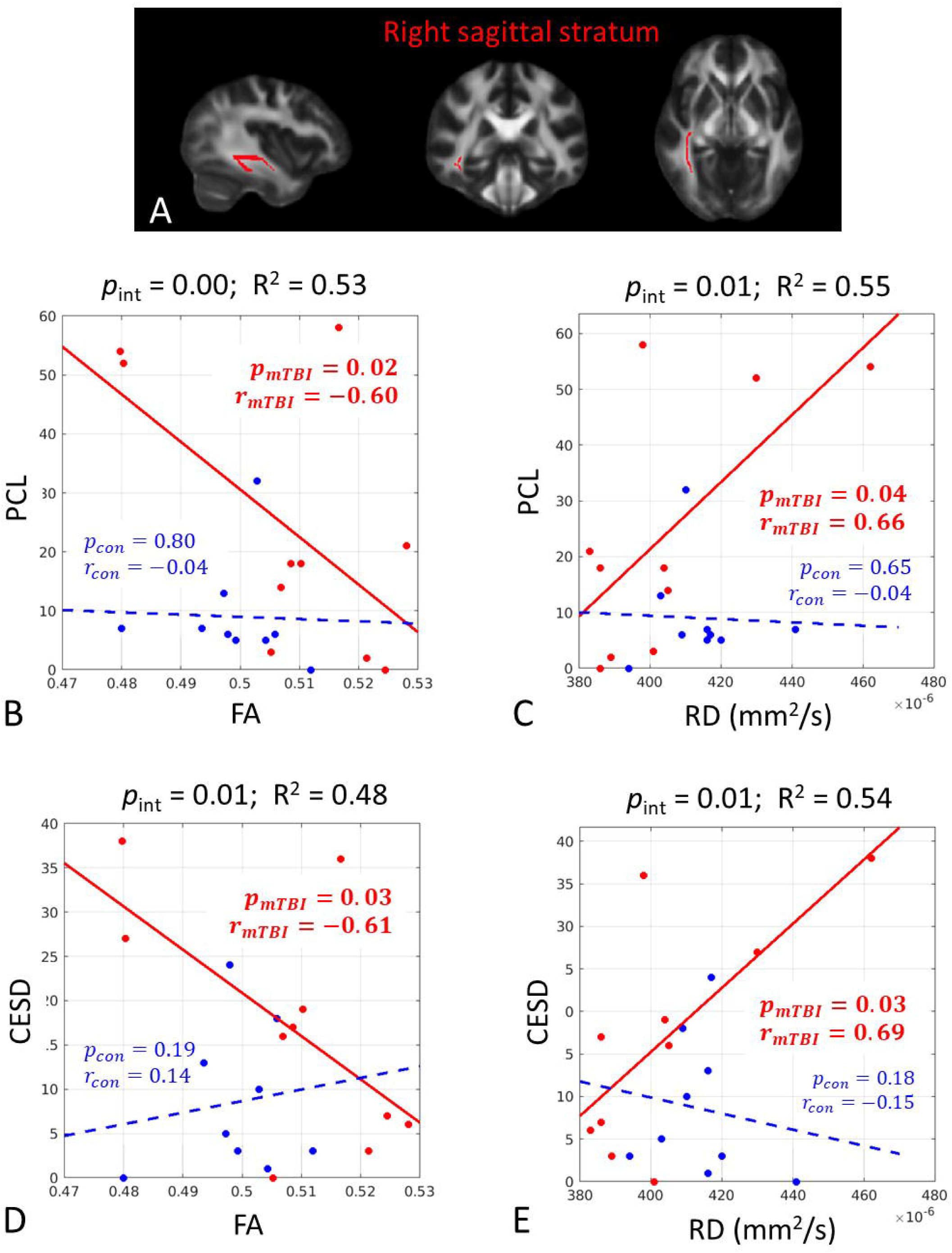
In the right sagittal stratum, there were significant group differences in the associations between the DTI metrics (FA and RD) at 1-month postinjury and the psychological measures (PCL and CESD) at 6-month postinjury. (A) The significant white matter tract is shown in red. (B-E) Scatter plots with post hoc correlation results. The mTBI group had a significant positive association (red solid line) while the control group did not (blue dashed line). Abbreviations: FA = fractional anisotropy; RD = radial diffusivity; PCL = posttraumatic stress symptoms questionnaire score; CESD = The depression scale; *p*_int_ = significance level of the group interaction term in Eq. 3; R^2^ = overall coefficient of determination for goodness of fit for Model 3; *p*_mTBI_ = significance level of the association for the mTBI group; r_mTBI_ = correlation coefficient for the mTBI group; *p*_con_ = significance level of the association for the control group; r_con_ = correlation coefficient for the control group Bold numbers indicate statistical significance with p < 0.05.

**Figure 5.**
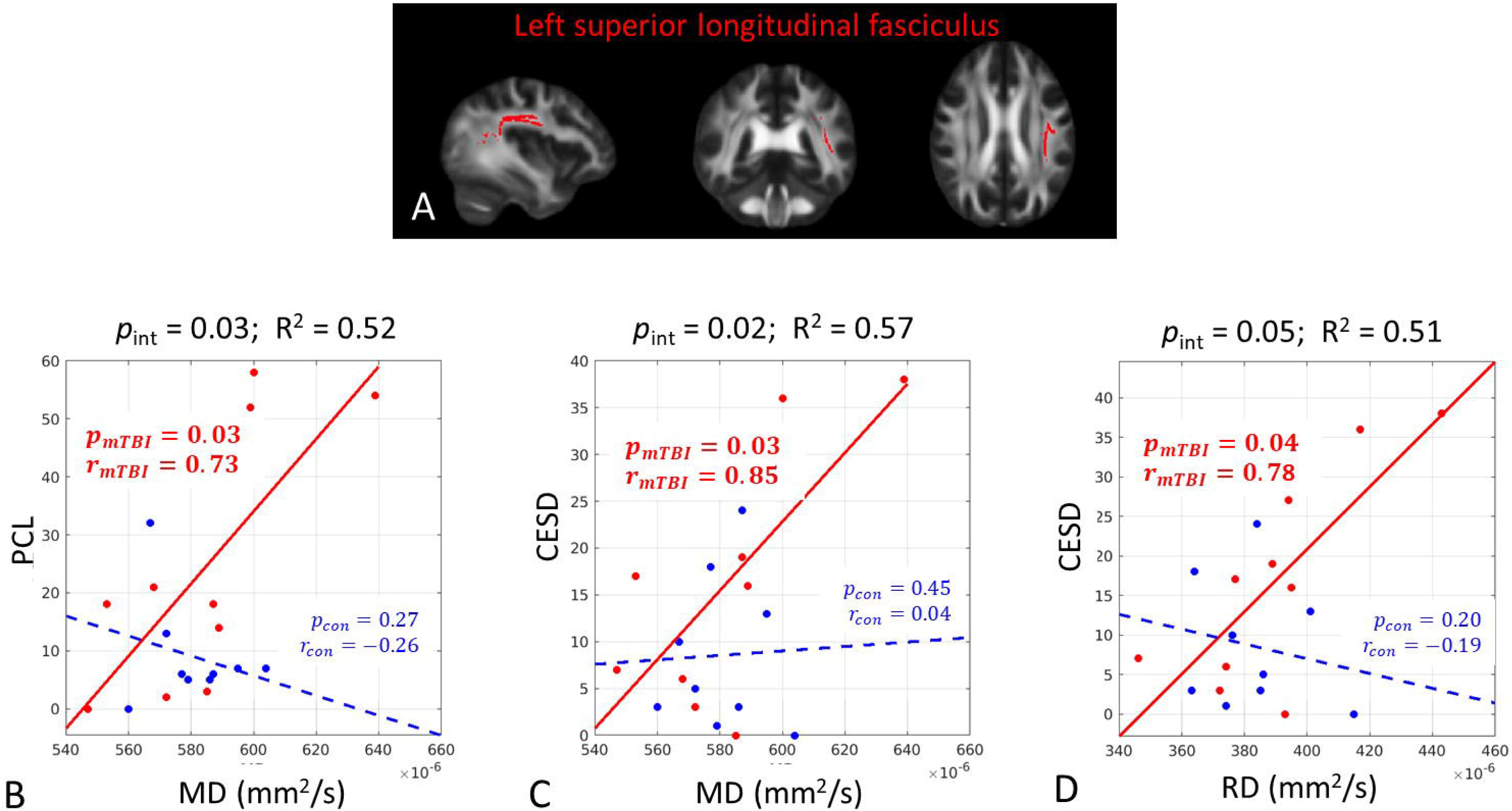
In the left superior longitudinal fasciculus, there were significant group differences in the associations between the DTI metrics (MA and RD) at 1-month postinjury and the psychological measures (PCL and CESD) at 6-month postinjury. (A) The significant white matter tract is shown in red. (B-E) Scatter plots with post hoc correlation results. The mTBI group had a significant positive association (red solid line) while the control group did not (blue dashed line). Abbreviations: MD = mean diffusivity; RD = radial diffusivity; PCL = posttraumatic stress symptoms questionnaire score; CESD = The depression scale; *p*_int_ = significance level of the group interaction term in Eq. 3; R^2^ = overall coefficient of determination for goodness of fit for Model 3; *p*_mTBI_ = significance level of the association for the mTBI group; r_mTBI_ = correlation coefficient for the mTBI group; *p*_con_ = significance level of the association for the control group; r_con_ = correlation coefficient for the control group Bold numbers indicate statistical significance with p < 0.05.

## DISCUSSION

This is the first study investigating association between white matter microstructure changes and Quantitative sensory tests (QST) (TS, PPT, CPM) in mTBI patients. In this study, we performed DTI imaging to detect white matter microstructural changes in mTBI and their relationships with the pain and psychological related measures. We did not find overwhelming group differences in the DTI metrics at 1-month postinjury across the whole cerebral white matter in this group of modest sample size (n = 12). We only observed reduced axial diffusivity in the forceps major in this subacute phase of mTBI. Previous studies have found significantly lower AD in mTBI participants in the left superior longitudinal fasciculus and the right anterior thalamic radiator, compared to healthy controls [47]. In addition, studies on migraine have found lower AD in forceps minor among migraineurs, as well as in the white matter fiber bundle connecting the mPFC and amygdala, which is associated with pain chronification [48, 49]. Pediatric migraineurs have also been found to have lower AD in the brain stem, bilateral thalamus, and fronto-temporo-occipital lobes, compared to healthy controls [50]. Additionally, previous studies also demonstrated significant DTI changes at 1-month postinjury with increased MD in the superior longitudinal fasciculus [51] and decrease MD at both left and right dorsolateral prefrontal cortex [52], whereas no change in DTI metrics were also reported [53].

Despite minimal absolute differences between group, we found that the early DTI metrics were predictive of later pain and psychological related measures. For the quantitative sensory tests, decreased white matter organization predicted increased endogenous facilitation of pain and decreased endogenous inhibition of pain. Additionally, decreased parallel water diffusion (i.e., axial diffusivity along the axonal orientation) predicted increased pain sensitization of the head area. The early DTI metrics were also sensitive to the psychological measures. Decreased white matter organization was associated with greater posttraumatic stress and depression symptoms. On the other hand, increased diffusivities were associated with later posttraumatic stress, pain catastrophizing scale and depression symptoms. Increased diffusivities were also associated with greater self-evaluated headache pain. The predictivity of the DTI metrics had significant group differences in the psychological measures and PPT, in which the control group did not exhibit such relationships.

For mTBI participants, overall directional changes in DTI metrics at 1-month postinjury associated with worse QST, psychological, and clinical measures at 6-month postinjury were decreased anisotropy (FA) and increased diffusivities (i.e., MD, AD, and RD). Such directionalities may indicate degraded white matter organizations with dispersed and impaired ordering of axonal fibers and increased intra– and extra-axonal space enabling freedom of water diffusion.

From our results, the most sensitive white matter tracts appeared to be the left insula, right sagittal stratum, and left superior longitudinal fasciculus. Insula is an essential component of the pain matrix for pain perception with extensive structural connections to the prefrontal, parietal, and central cingulate gyri [54–56]. Striatum, composed by caudate, putamen, and ventral striatum, receives afferents from cortex, midbrain and thalamus and deliverers signal to the basal ganglia [57, 58]. The neural activity in striatum is mainly response to movements and rewards [59, 60]. The superior longitudinal fasciculus (SLF), which connects the frontal, parietal, and occipital lobes [47], is highly relevant to prefrontal cortical pain modulation [61, 62].

Our results are supported by previous studies. A previous study showed mTBI patients with poor outcomes on clinical tests had significant MD increase in bilateral superior longitudinal faciculus three to four months after injury [63]. Additionally, significant positive associations were found between headache frequency and bilateral cingulum angular bundles MD in persistent PTH patients and between headache frequency and forceps major MD in migraine patients [44]. Apart from diffusion findings, a previous study reported that the gray matter volume (GMV) decreased in the insula and increased in the ventral striatum for patients with medication-overuse headache [64]. The decreasing GMV in right insula was also found in mTBI patients within 7 days of injury [65].

In the current study, the clinical pain and psychological measures were more sensitive than the quantitative sensory tests with respect to group differences and relationships with DTI. Several explanations could underlie these findings. For example, the DTI metrics were predictive of headache pain and the psychological measures in mTBI, but not controls. This lack of relationship in the controls could be due to the overall lower scores and variability in the clinical pain and psychological measures within the control group. Regarding the QST measures, our group has shown in a prior study with a larger sample size that mTBI patients exhibit abnormal pain modulation on the QST tests in the acute and subacute stages of injury that appear to normalize into chronic stage of injury [66]. These findings could help explain why the relationships between the DTI metrics and QST measures of pain modulation at 6 months post injury did not differ between the mTBI and control groups.

There are several limitations in this study. The primary constrain was the modest sample sizes, which minimized the capability of detecting small effect sizes. Also, as a preliminary study, this study could not afford adjustments for multiple comparisons. Nevertheless, with multi-steps of direct association analyses and interaction analyses, our results can still provide the community useful knowledge and effect size for future large scale study designs. Another limitation stems from the diffusion tensor assumption, in which Gaussian distribution is used to summarize overall water diffusion behavior within an imaging voxel comprising multiple tissue types, crossing fibers, and diffusion compartments. We had limited our analyses in the center of the white matter tracts (skeletonized white matter ROIs), where the diffusion tensor assumption is less violated. Future diffusion MRI could focus on biophysical modeling, such as neurite orientation dispersion and density imaging [67] or kurtosis-based white matter tract integrity imaging [68], to produce more biologically specific diffusion metrics.

## Data Availability

De-identified data from this study will be made publicly available in (FITBIR)

## Acknowledgements

Data presented herein were obtained at the Department of Radiology and Imaging Sciences at Indiana University School of Medicine.

## Authors’ contributions

FAW, KMN, and YCW conceptualized the study and designed the experiments. HCY and TN carried out the experiments. HCY and KMN analyzed the data. YCW wrote the manuscript, with guidance from FAW and KMN. All authors read and approved the final manuscript.

